# Impacts of COVID-19 public measures on country-level trade flows: Global panel regression analysis

**DOI:** 10.1101/2021.07.09.21260294

**Authors:** Sylvia Szabo, Worawat Srisawasdi, Takuji W Tsusaka, Reuben M.J. Kadigi, James Vause, Neil D. Burgess

## Abstract

As of 10 July 2021, there have been over 186 million cases of COVID-19 and more than four million died as a result of this disease. The COVID-19 outbreak has also contributed to tremendous global decline in trade flows. The rapid spread of COVID-19 and the measures implemented by governments to contain the pandemic have had serious consequences for the world’s economies. While the pandemic has affected the international movement of people, goods and services, there is still limited systematic research regarding the possible associations between the COVID-19 measures on countries’ international trade flows. To fill this gap, we conducted regression analysis based on country level time series data from the United Nations and World Bank datasets. The results of the random effects panel regression models show that, the country import and export values are positively affected by health-related policies, while there is a negative association between stringency measures and import and export values. More specifically, school closing, stay-at-home requirements, and testing policy measures were found to have significant negative effects on countries’ trade values. In contrast, facial covering policies were found to have significant positive effects on countries’ import, export and total trade values.

## 1. Introduction

Since the outbreak of COVID-19, more than four million people have lost their lives due to the pandemic (WHO 2021). The number of new COVID-19 cases has been rising at an alarming rate and many countries have already experienced several ‘pandemic waves’ (Diaz 2020). While uncertainty still remains as to how and when the pandemic will run its course, the unprecedented economic impact caused by the global health emergency has already sharply exposed the global economy’s existing weaknesses in complex supply chains, and is severely slowing down the development progress around the world (UNCTAD 2020). Data show that more people are living in extreme poverty as the global economy experienced sharp contraction in 2020 (UNCTAD 2020).

Recent literature shows that the COVID-19 pandemic has had a direct impact on global economy in a number of ways. Literature surveys by Goodell (2020) and Yarovaya et al. (2020) further showed that the COVID-19 pandemic has had a direct negative impact on the financial sector. Bachman (2020) pointed out that the COVID-19 pandemic has negatively affected global production, disrupted supply chains and unsettled financial markets globally. Ashraf (2020b) found that higher national-level uncertainty significantly strengthens the negative stock markets’ reaction to an increase in COVID-19 confirmed cases. The decline in purchasing power linked to lost income due to COVID-19 pandemic has threatened food security, disrupted the global supply chain, and interrupted the movement of migrant workers (UNEP 2020).

Some past literature has explored COVID-19’s impact on globalization. Shrestha (2020b) examined the potential impact of COVID-19 on globalization and global health in terms of mobility, trade, travel, and countries most impacted. The results showed that the pandemic has affected the world economy, healthcare, and globalization through travel, events cancellation, employment workforce, food chain, academia, and healthcare capacity. Certain countries were more vulnerable than others. In Africa, more vulnerable countries included South Africa and Egypt. In Europe, Russia, Germany, and Italy, and in Asia and Oceania, India, Iran, Pakistan, Saudi Arabia, and Turkey. For the Americas, vulnerable countries include Brazil, USA, Chile, Mexico, and Peru (Shrestha et al. 2020b).

The COVID-19 outbreak has led to a contraction in the volume of international trade. For the whole of 2020, trade volume was down 5.3% compared to 2019 (WTO 2021). Exports from large economies including the United States of America (USA), Japan, and the European Union (EU) have been particularly affected. It should be noted that the economic contraction in China was smaller than the global average, as China controlled the outbreak and reopened its economy relatively quickly. At the time of writing, Latin America and the Caribbean were the developing regions which were most affected by the pandemic. Manufacturing activities have been disrupted, first in Asia and then in Europe, North America and the rest of the world (UNECLEC 2020). Widespread border closures have resulted in a steep rise in unemployment, especially in USA, which led to a reduction in the demand for goods and services. Global GDP in 2020 registered its sharpest contraction since the Second World War (UNECLAC 2020).

The rapid spread of COVID-19 and the public measures implemented by governments to contain it have had serious consequences for the world’s economies and trade flows. Maliszewska (2020) simulated a model showing the potential impact of COVID-19 on GDP and trade, using a standard global computable general equilibrium model. The results indicate a fall of world’s GDP by 2% below the benchmark for the baseline global pandemic scenario and the declines are nearly 4% for amplified pandemic scenario. The biggest negative shock was found in the following areas: reduction in the domestic services output, an increase in international trade costs, a drop in travel services, and reduction in activities that require proximity between people. The World Bank projects that as a result of the COVID-19 crisis the global output will fall between 5.2 and nearly 8 per cent in 2020 and the World Trade Organization projects global merchandise trade to fall by between 13 and 32 per cent in optimistic and pessimistic scenarios (WB 2020; WTO 2020a). Countries were affected by sharp decline in exports, particularly to the European Union and the United States.

The COVID-19 pandemic has also sparked concerns regarding export restrictions being imposed by countries around the world during the pandemic. In an effort to control the pandemic, some countries decided to establish export controls over certain medical products in the form of temporary export bans or due to additional requirements for licensing and authorization. Other countries, concerned with the sufficiency of food supplies, have introduced export restrictions over agricultural products. These decisions have implications for equitable food distribution in the global market (Nguyen 2020).

While there is already emerging literature on the impact of COVID-19 on some aspects of the economy (Ashraf 2020b), there is still no systematic quantitative examination on the effects of pandemic-related policies and measures on global trade flows. Our paper aims to fill this gap by investigating the impacts of the COVID-19 government response measures on export and imports as well as overall trade flows. In other words, we set out to identify the policy clusters as well as specific government policies and measures that have significant impacts on import and export values in 2020 amid the COVID-19 pandemic. By undertaking this analysis, we aim to contribute not only to the existing literature but also to the ongoing policy discussions and respective choice of policy instruments aimed at effectively controlling the spread of COVID-19 and maintaining positive economic growth.

## 2. Hypothesis testing

In this section, we present three testable hypotheses regarding the direct and indirect impact of government social distancing measures, containment and health policies and economic support programs to test the impact of COVID-19 government response measures on trade flows. The three government response indices include stringency index (SI), containment and health index (CHI) and economic support index (ESI). CHI, which represents the government emergency policies, is constructed based on from 3 specific policies, i.e. public awareness campaigns, testing policy and contact tracing. SI records data on social distancing measures. The stringency policies include school closing, workplace closing, cancel public events, gathering restrictions, public transport closing, stay at home requirements, and restrictions on internal and international travel. ESI is composed from 2 policies including the government income support and debt relief for households. These economic support policies aim to provide income support to citizens amid crisis (Hale et al. 2020b).

**Hypothesis 1**: Health-related policies have a positive impact on trade flows.

The CHI include three specific policies, i.e. public awareness campaigns, testing policy and contact tracing (Ashraf 2020b). Our hypothesis is that government containment and health policies have positive effects on trade flows. Ashraf (2020b) shows that the government containment and health policies have a significant negative effects on stock market returns. The government intensive information campaign provides awareness about the benefits of staying at home, sanitization, frequent testing and contact tracing can help to identify infected and suspected cases early on and reduce the number of infected cases (CDC 2020). Such effective measures can boost the public’s confidence in the government’s ability to deal with the pandemic as well as effectively control the spread of COVID-19 viruses. Consequently, good management of the pandemic including more lives saved, and reduced overall impacts can support overall economic performance (Greenstone and Nigam, 2020; Thunström et al., 2020). The government pandemic management measures can help strengthen economic activities and increase the trade volume during the pandemic crisis.

**Hypothesis 2**: Closure and social distancing measures are likely to decrease trade value.

Our assumption is that government closure and social distancing measures will reduce trade activities. However, despite the pre-supposed direct negative effect on trade activities, social distancing might have some positive economic impact through reducing the risk of COVID-19 death rates (Thunström et al. 2020). According to Greenstone and Nigam (2020), moderate social distancing in the USA could save up to 1.7 million lives. The net benefit of social distancing in the USA is estimated at $5.2 trillion (Thunstr et al. 2020). Countries where government implemented stringent social distancing policies are more likely to have their people complying with these policies and hence have higher chances of limiting the spread of the virus (Hussain 2020). This in turn can have a positive effect on the overall trade flows.

**Hypothesis 3**: Economic support policies lead to an increase in trade value.

Economic support programs may have positive effects on trade flows due to reduced infection rates that stem from enhanced capacity to comply with restriction measures which in turn leads to a healthier population able to produce more for own consumption and more surplus for sale to earn more income. Some studies found correlations between stay-at-home orders and income. In fact, lower income people may be less likely to comply with government restrictions, thus they are at a higher risk of getting infected (Lou et al. 2020). This points to a pressure experienced by lower income households to leave the house for income-seeking activities. Generous income support programs may thus help control the COVID-19 infection rates by allowing the lower income individuals to be able to stay at home (Wright et al. 2020).

## 3. Materials and methods

### 3.1 Data collection

We collected the data for this study from three major sources and categorized variables roughly into three groups, i.e. trade, COVID-19 new cases, and data related to public measures implemented by governments. Table 1 summarizes a list of 70 sample countries for which we were able to obtain the data. This table includes a summary of trade values of a second quarter for the period 2019 to 2020, total cumulative COVID-19 cases, as well as the Government Response Index (GRI) by country. First, we obtained the trade data, more specifically the values of imports, exports and total trade, from the UN Trade Statistics Database (UN Comtrade, 2021). The data used in this study is considered a panel data recorded chronologically on a monthly basis. The period of data collected stretches from January to August 2020 across the 70 sample countries. Second, we downloaded the data of daily COVID-19 confirmed cases for each country from the World Health Organization database (WHO, 2020). Third, we collected the data of COVID-19 government response indices and policies from the Blavatnik School of Government, University of Oxford (Hale et al., 2020b).

**Table 1:**
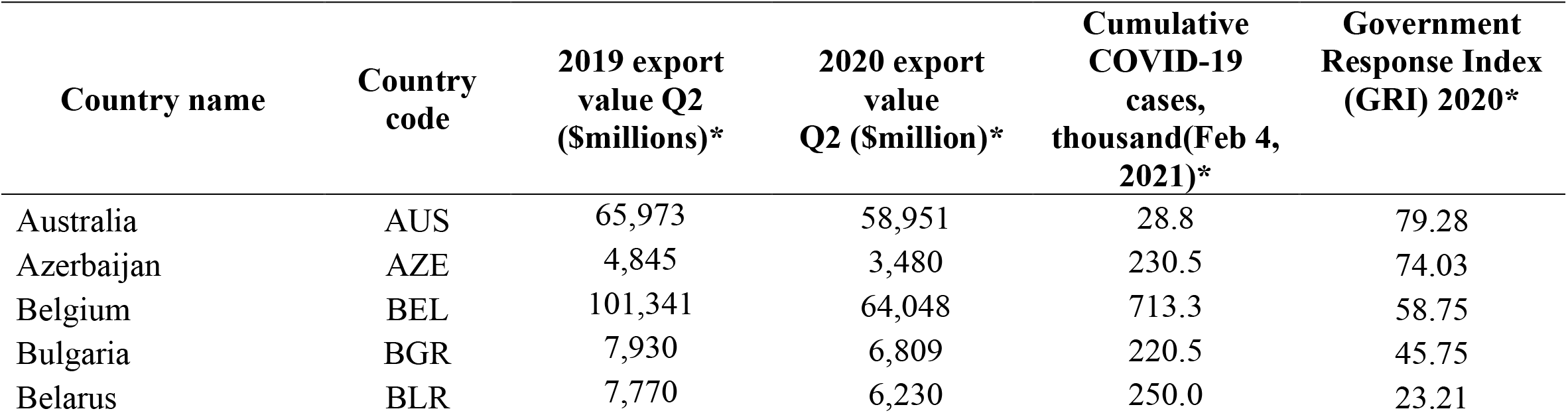
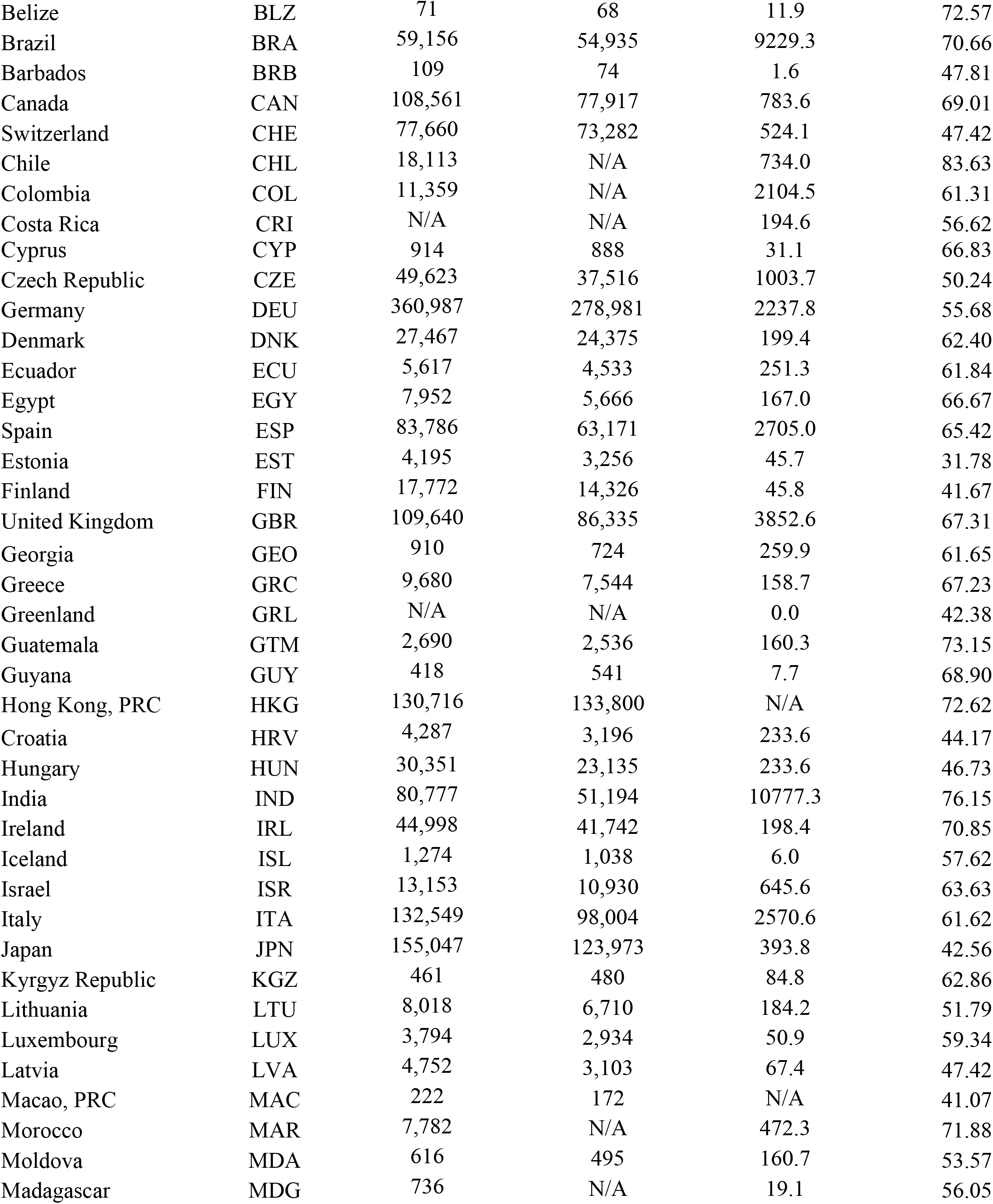
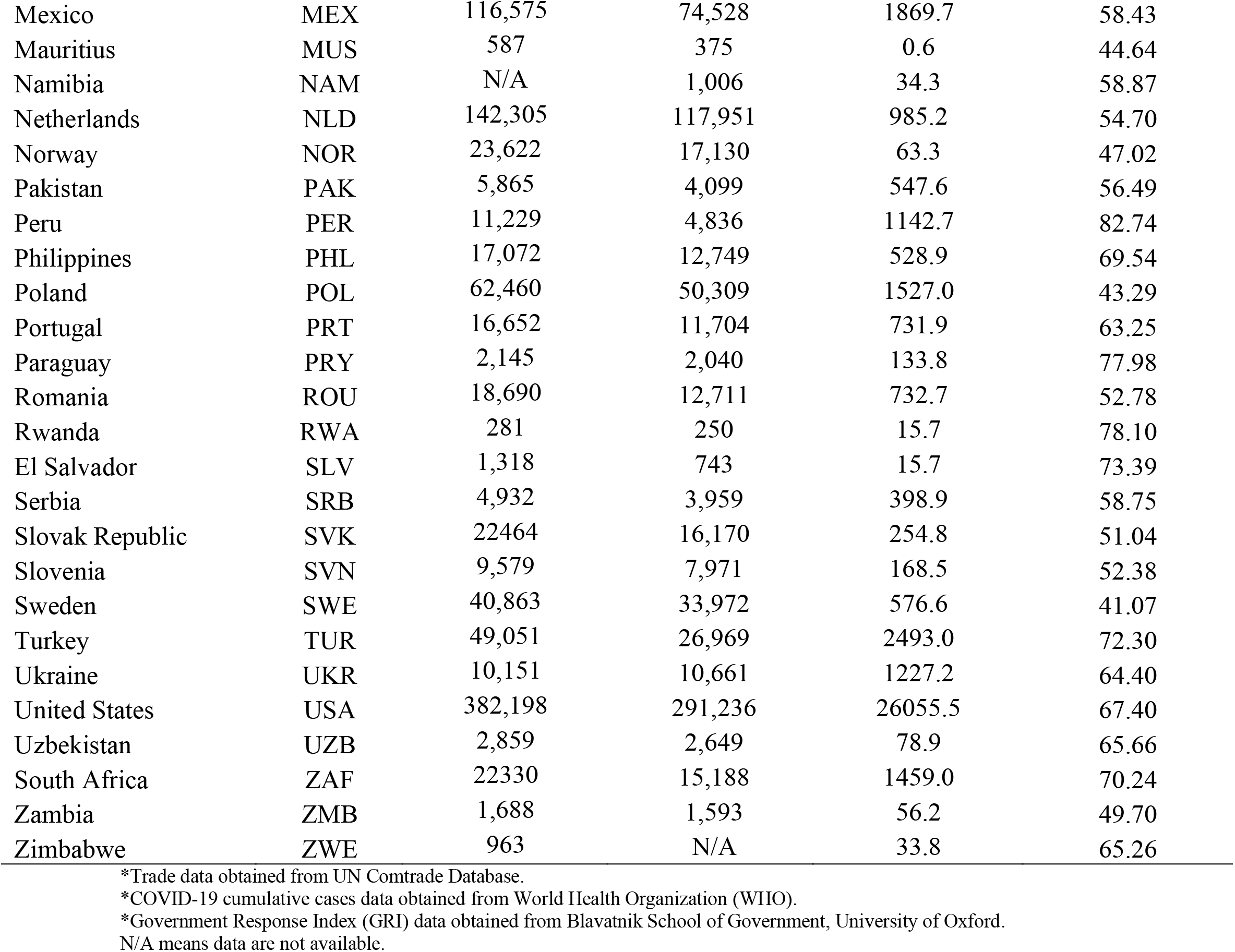
List of 70 sample countries with export values from 2019-2020(Q2), cumulative COVID-19 cumulative cases, and Government Response Index (GRI) by country

**Table 2:**
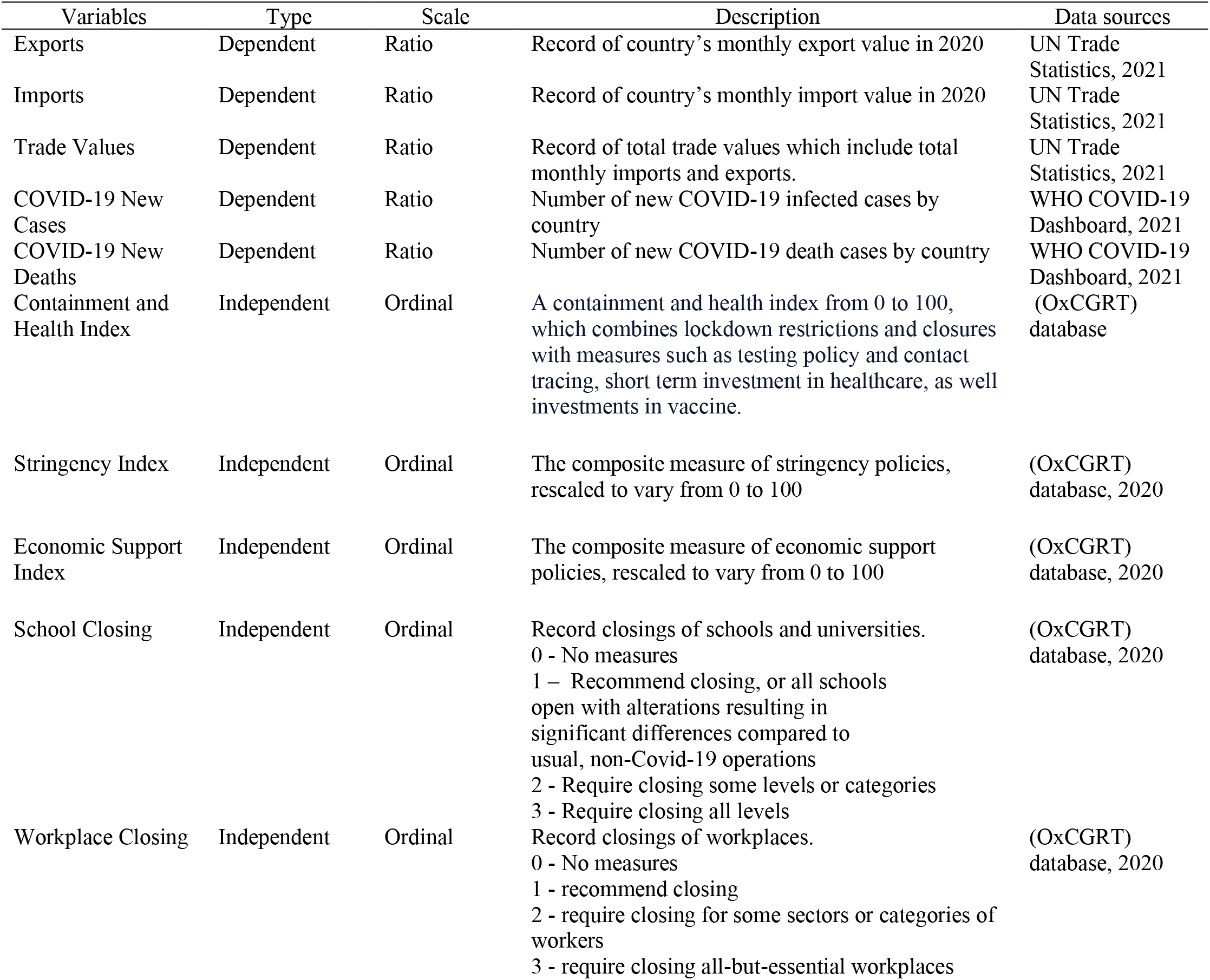
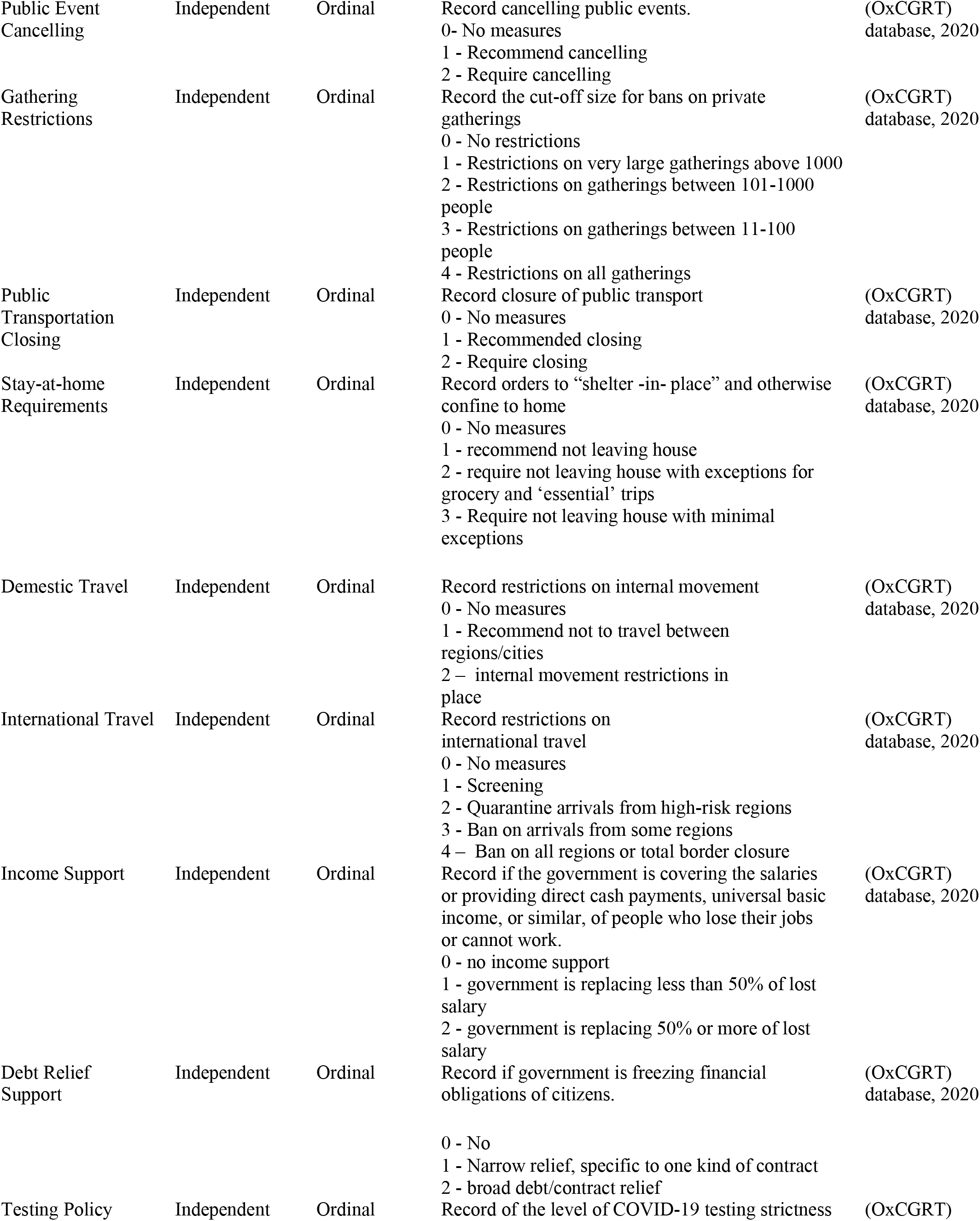
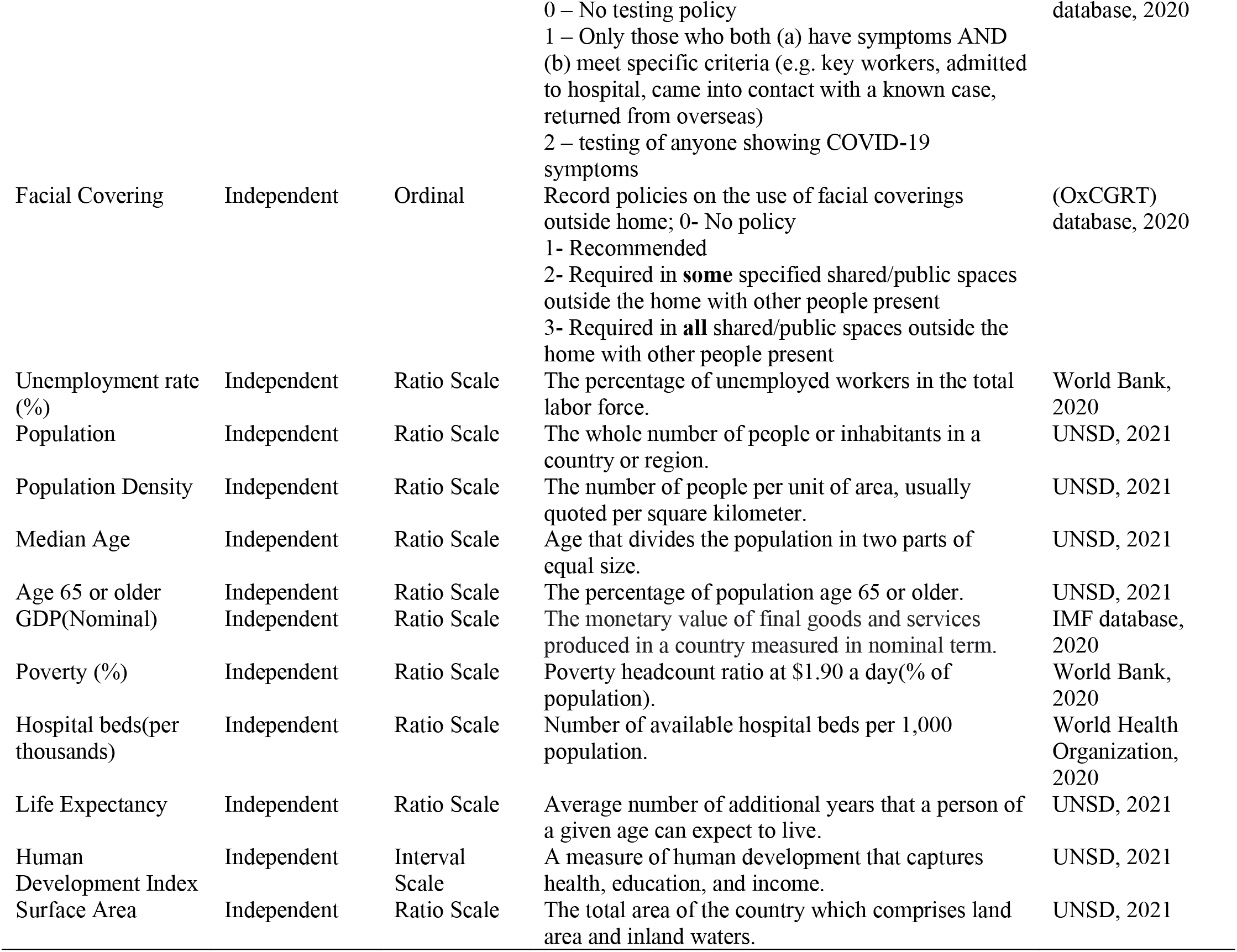
Variables Definition

Figure 1 shows a comparison of export values of the 2^nd^ quarter in 2019 and 2020. The data in this chart consists of information from 70 sample countries that we used in this study. The bar on the left side (*“2019 export value Q2”)* represents export values of 2019 and the bar on the right (“*2020 export value Q2”*) represents 2020 export values. As can be noted, export values declined significantly in 2020, compared to the same period (2nd quarter) in 2019. We chose the second quarter of 2020 to represent the change in export values because the COVID-19 pandemic only became a global threat in the very end of March 2020. Thus, before the effects of COVID-19 pandemic did not seriously hinder major economic activities around the world to an extent that would allow for the observation of a major decline. In the second quarter of 2020, however, the effects of COVID-19 pandemic started to show serious negative impacts on economic activities, including international trade. Most countries started to experience a significant decline in trade volume in 2020 from Q2, with the exception of some small low-income countries whose primary source of income does not derive from international trade.

**Figure 1:**
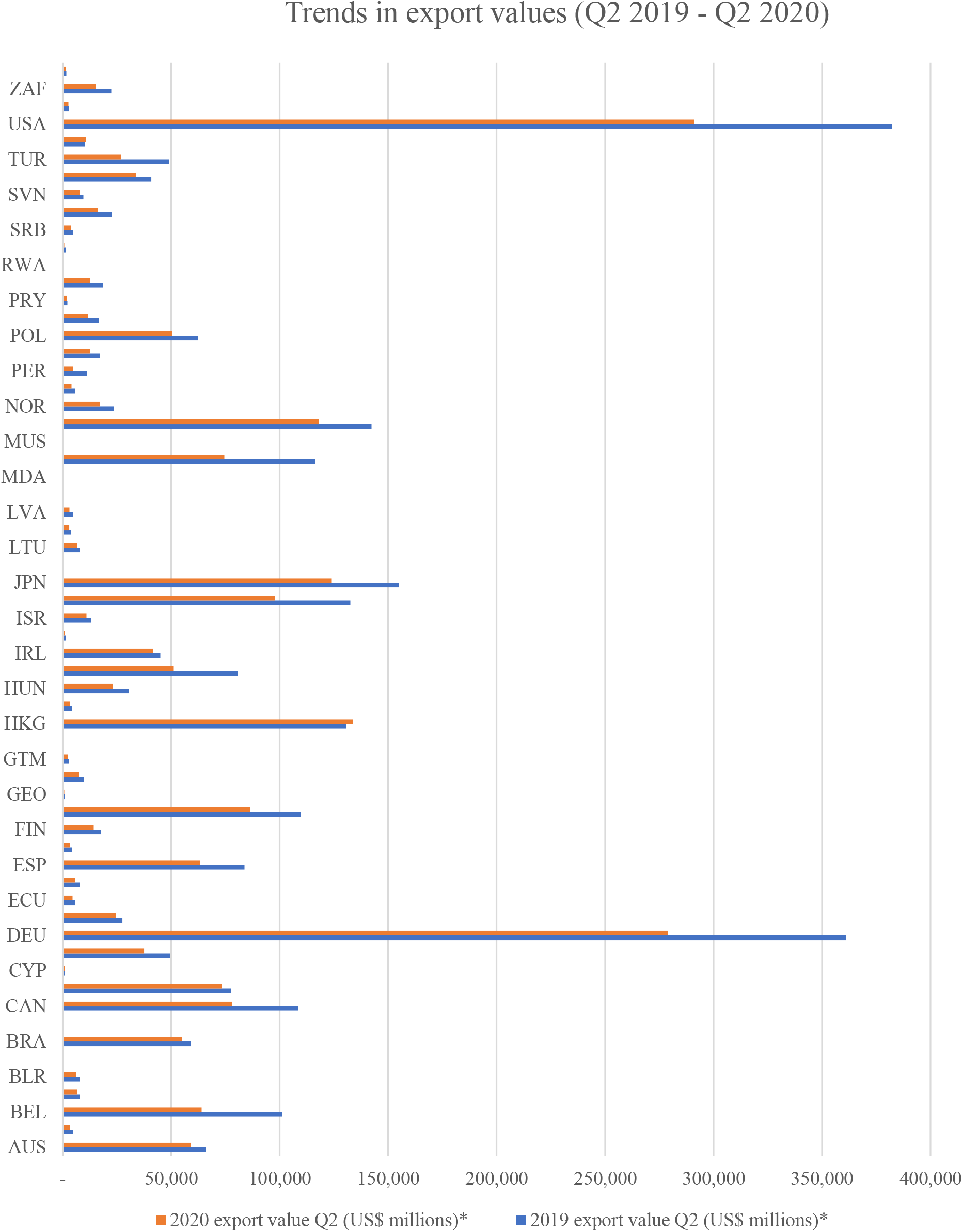
Export value comparison between the second quarter of 2019 and 2020 showing the impact of COVID-19 on international trade.

Figures 2 and 3 show gradual monthly changes in government intervention indices and specific measures indices from January to September 2020. The government intervention indices include GRI, CHI, SI and ESI. The specific measures in figure 3 include school and workplace closing, public event cancelling, gathering restrictions, public transportation closing, stay at home requirements, domestic and international travel restrictions, income support, debt relief, public awareness campaign, testing policy and facial covering. The values of government intervention indices (Fig. 2) were lowest in January and February 2020. The values gradually increase from March 2020 and peaked during April and May. Figure 3 shows that there was a gradual increase in specific policy indices. Restriction measures on limitation of traveling and public gatherings remain relatively high through March to September 2020. Other specific measures also show sharp increase from March onward. The values of specific indices reached the highest point during April and May. It can be inferred that since the COVID-19 pandemic started to spread globally from March 2020 onward, the stringency in the implementation of government intervention measures has remained high for the remainder of the year.

**Figure 2:**
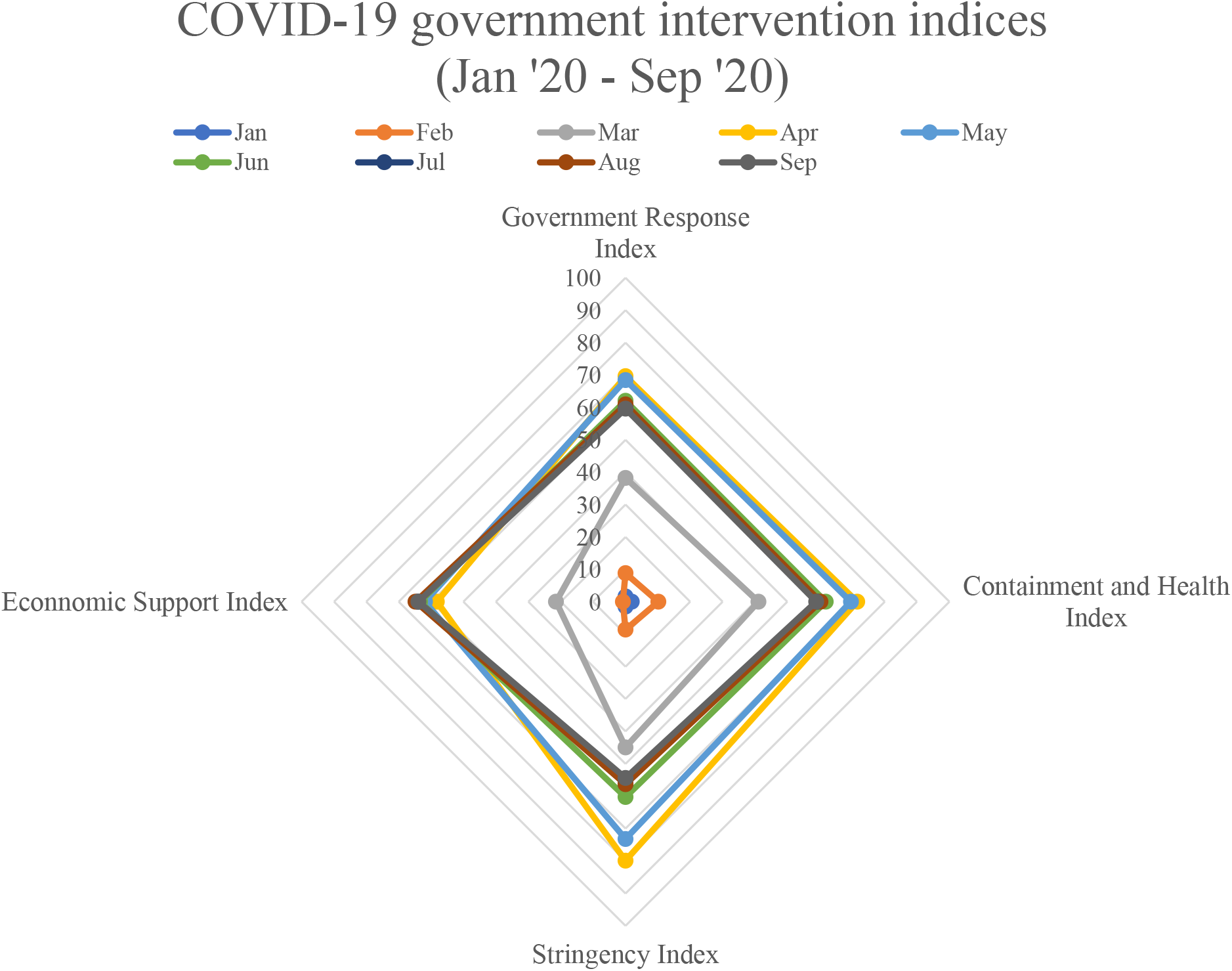
Government intervention indices (Government Response Index, Containment and Health Index, Stringency Index and Economic Support Index) from January 2020 to September 2020.

**Figure 3:**
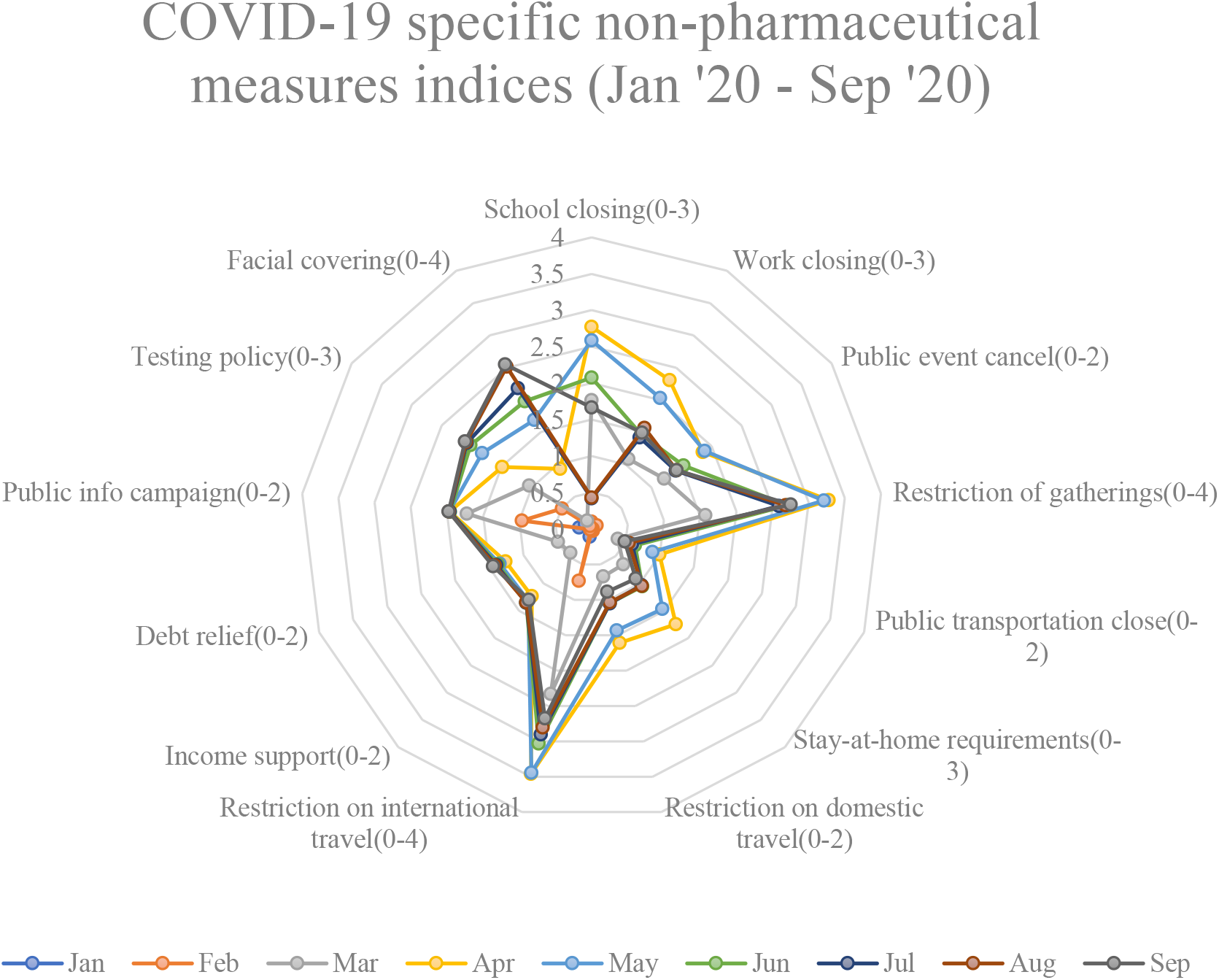
Monthly changes in COVID-19 government specific measures from January 2020 to September 2020.

Figure 4 and 5 show the gradual increase in the number of COVID-19 daily new cases and deaths in six different regions (Africa, Asia, Europe, Oceania, North America, and South America) from January to September 2020. More specifically, there was a sharp spike in number of daily cases from March 2020 onward (Fig. 4). Europe and North America in particular experienced a sharp increase in daily cases compared to other regions during April. From May 2020 onward, other regions including Asia and South America also faced sharp increases in daily cases. Africa and Oceania appeared to be the least affected regions. The number of daily cases in Oceania region in particular remained relatively low throughout the year compared to other regions. In contrast, Asia, Europe and North America were shown to be heavily affected by the substantial increase in daily cases throughout March to September. It can also be observed that the daily new deaths show similar overall trend compared to daily new cases (Fig. 5). However, South Africa appeared to experience high fluctuation in death cases, reaching over 5,000 daily deaths in certain periods. This could be due to poor management of healthcare system compared to other regions. Europe on the other hand tend to experience gradual decline in death cases. Official reported daily death cases in Oceania and Africa remain lower compared to other regions due to having less number of infected cases. Death cases in Asia and North America remain relatively high throughout September 2020.

**Figure 4:**
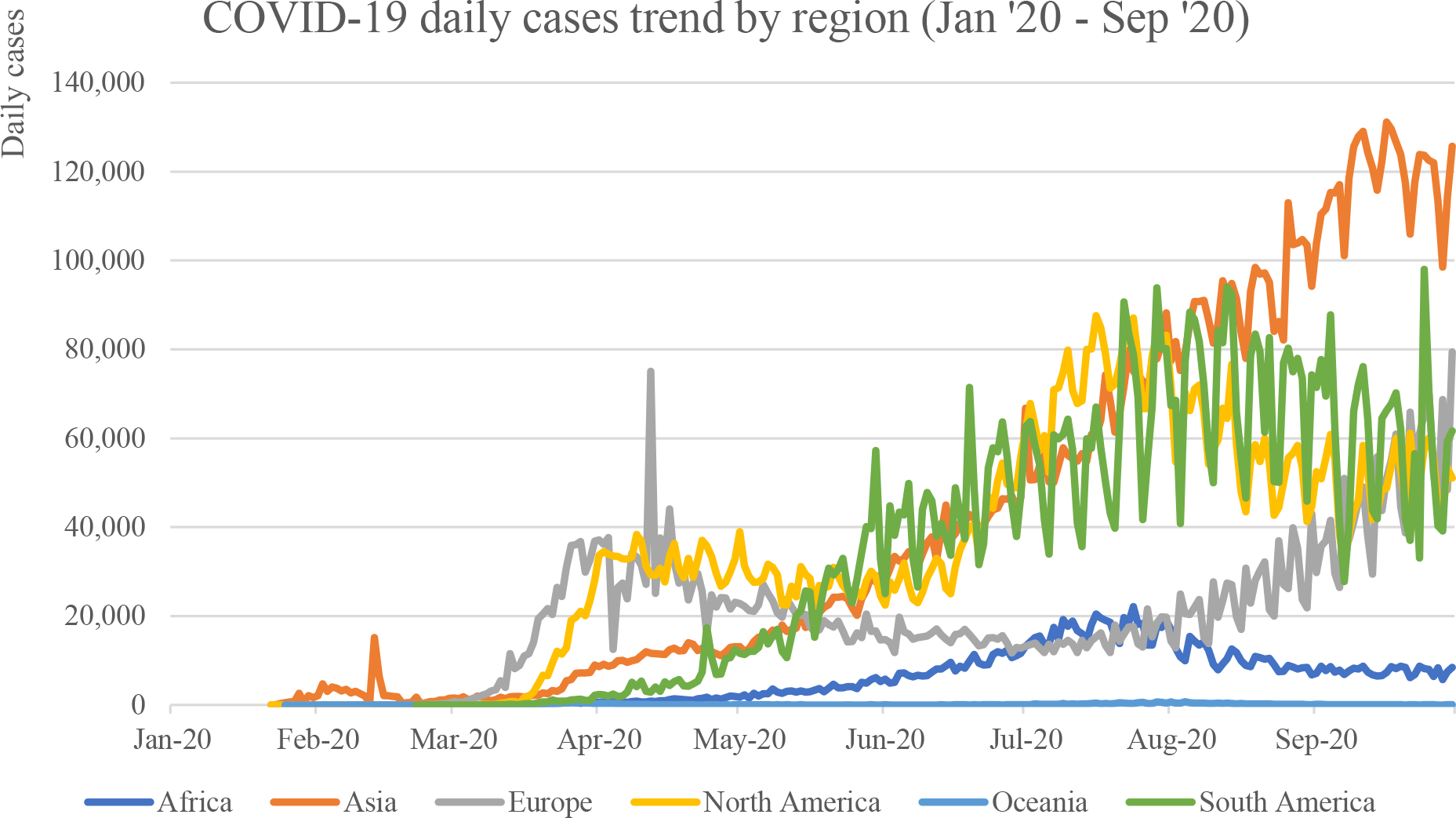
COVID-19 daily cases by region from January 2020 to September 2020.

**Figure 5:**
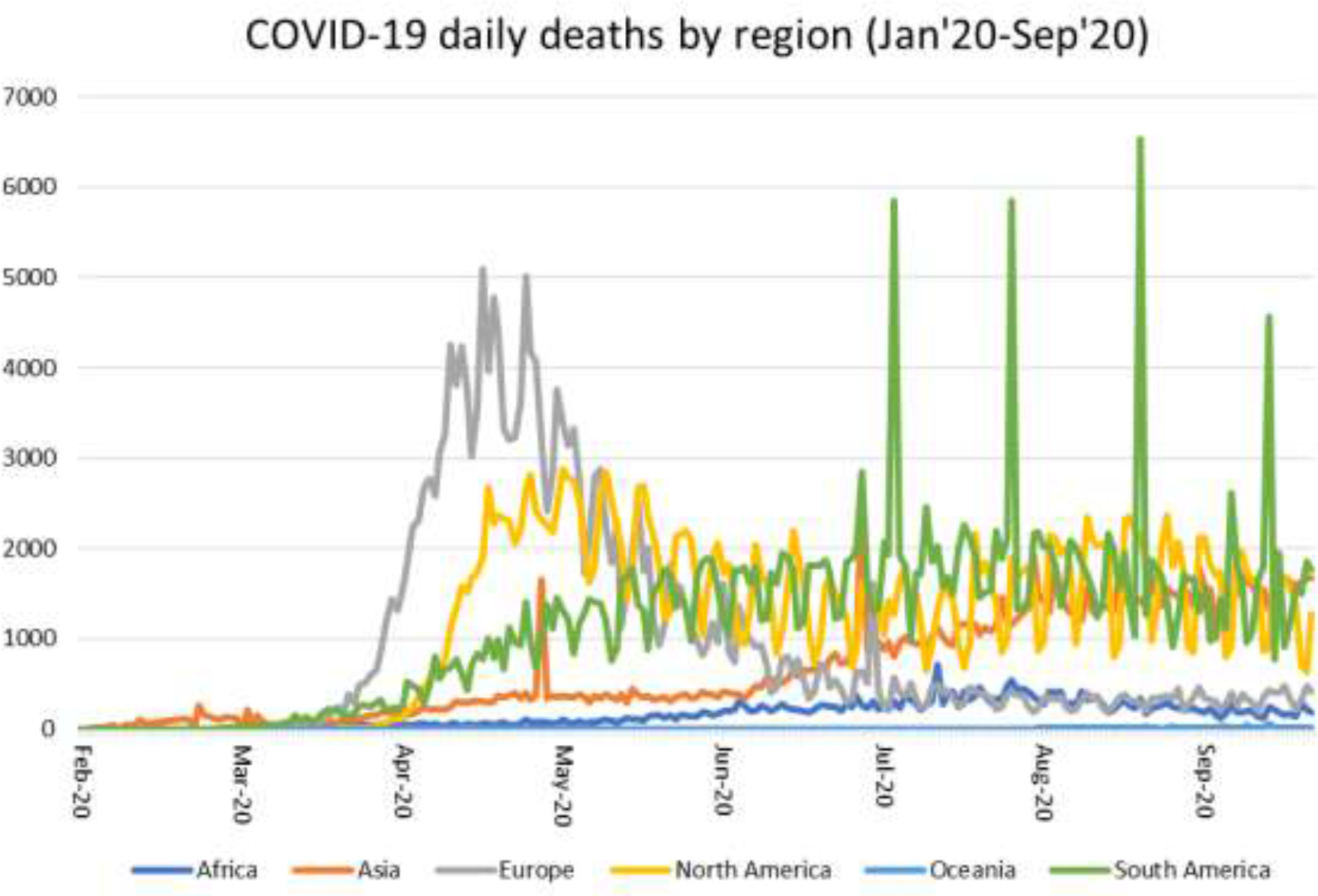
COVID-19 daily new deaths by region from January 2020 to September 2020.

Figure 6 illustrates the effective reproduction rate (R) of regions around the world from January to September 2020. Effective reproduction number is defined as the average number of secondary cases produced by a primary case (Arrryo-Marioli et al. 2021). The effective reproduction number remained high during February to March 2020. The effective reproduction number gradually declined to around 1.0-1.5 from April 2020 onward. In Oceania region, effective reproduction number remained relatively low compared to other regions. This is supported by the fact that the number of daily cases for Oceania region remained relatively low compared to the rest of the world. On average, effective reproduction number for other regions were above 1.0. In this context, the number of infected individuals will keep increasing as long as *R* > 1. Therefore, it can be assumed that other regions apart from Oceania were still facing increase in number of infected individuals throughout 2020.

**Figure 6:**
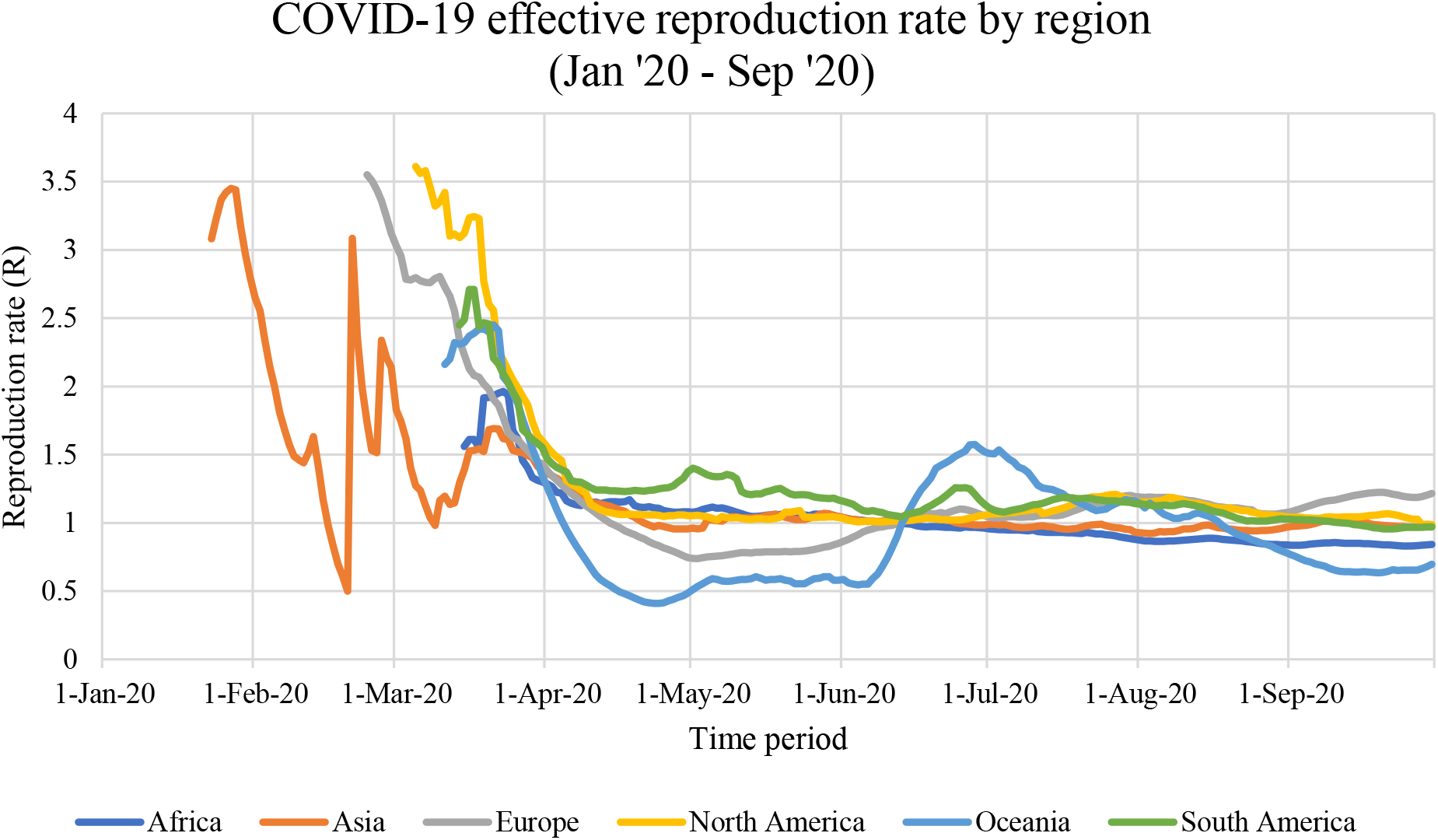
COVID-19 effective reproduction number by region from January 2020 to September 2020.

### 3.2 Measurement of trade flows and government response policies

In this study, the country’s imports, exports and total trade values are assigned as dependent variables. The independent variables include government policy indices and government specific policies and control variables. The government policy indices include 3 distinct indices; containment and health index, stringency index and economic support index (Hale et al. 2020b). The indices are simple averages of the individual component indicators. The government specific policies include specific indicators measured in ordinal scale, which include school closing, workplace closing, cancel public events, gathering restrictions, stay at home requirements, domestic movement restriction, international travel restriction, income support, debt relief, public info campaign, testing policy and facial coverings. The control variables include country’s geographic, demographic and economic variables such as GDP, population, population density, life expectancy and surface area.

For government policy indices, we used the Oxford COVID-19 Government Response Tracker database (Hale et al. 2020b). As mentioned previously, there are three main indexes in this database: CHI, SI and ESI. SI records information on closure and social distancing measures using 8 indicators including school closing, workplace closing, cancel public events, restrictions on gathering size, close public transport, stay at home requirements, restrictions on internal movement and restrictions on international travel. ESI is based on 2 specific policies, including the government income support and debt/contract relief for household programs. CHI was coded from indicators for containment policies as well as health system policies, which include public information campaign, testing policy, contact tracing, and facial covering policy (Hale et al. 2020b). It represents government policies concerning healthcare system in the country. Each of the three indices was calculated and rescaled to have a range from 0 to 100. For specific policy variables, we chose the individual policy indicators that make up the containment and health, stringency and economic support indices.

### 3.3 Statistical methods

The random effects regression modelling was used as a method for the analysis. Random effects models are applicable in the context of analyzing data with characteristics that fit the description of panel data (Arellano 2003). That is, data comprising observation units on two or more groups over two or more time periods. In this case, the data were from 70 countries observed over 9 monthly periods, which fits the panel data description.

In econometrics, the random effects models are used in panel data analysis when one assumes no fixed effects (it allows for individual effects). The random effects model is a special case of the fixed effects model. Random effect models assist in controlling for unobserved heterogeneity when it is constant over time and not correlated with independent variables. This constant can be removed from longitudinal data through differencing, since taking a first difference will remove any time invariant components of the model (Woodridge 2010). The rationale behind random effects model is that, unlike the fixed effects model, the variation across entities is assumed to be random and uncorrelated with the predictor or independent variables included in the model (Green 2008).

An advantage of random effects is that it allows for including time invariant variables (i.e. gender). In the fixed effects model these variables are absorbed by the intercept. The random effects model can be presented as in Equation 1:

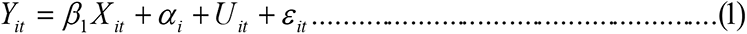

where

*α*_*i*_ is the unknown intercept for each entity (n-entity-specific intercepts)

*Y*_*it*_ is the dependent variable (DV) where i = entity and t = time,

*X*_*it*_ represents one independent variable (IV),

*β*_1_ is the coefficient for that independent variable,

*U*_*it*_ is the between-entity error term,

*ε*_*it*_ is the within-entity error term

In this paper, we introduce three models for analyzing the impacts of COVID-19 related public measures on trade. As mentioned previously, the three policy indices include health and containment, stringency and economic support indices. In addition, we introduce another model to identify the impacts of government specific policies on trade during the COVID-19 pandemic. The components of government response indices included as independent variables are CHI, SI and ESI. Furthermore, we include specific policies variables for analyzing the impact of specific policies on trade. The set of specific policies include school closing, work closing, public event canceling, gathering restrictions, stay-at-home requirements, domestic and international travel restrictions, income support, public campaign, testing policy, debt relief, and facial covering. In addition, variables such as country’s demographics and geographic and economic variables are added into the model to increase the robustness of the model’s results.

## 4. Results

Table 3 shows a summary of descriptive statistics generated from the data we collected in this study. The descriptive summary illustrates the mean, standard deviation, minimum and maximum values of the data collected from 70 sample countries. To avoid collinearity issues between independent variables, we use the Variance Inflation Factor (VIF) analysis to identify and remove independent variables with high VIF values. The dependent variables in this study include the monthly exports, imports and total trade values. The independent variables include COVID-19 government policy indices and specific policies and actions. Furthermore, we also include control variables to improve robustness of the model such as country’s economic, demographic, and geographic variables.

**Table 3:**
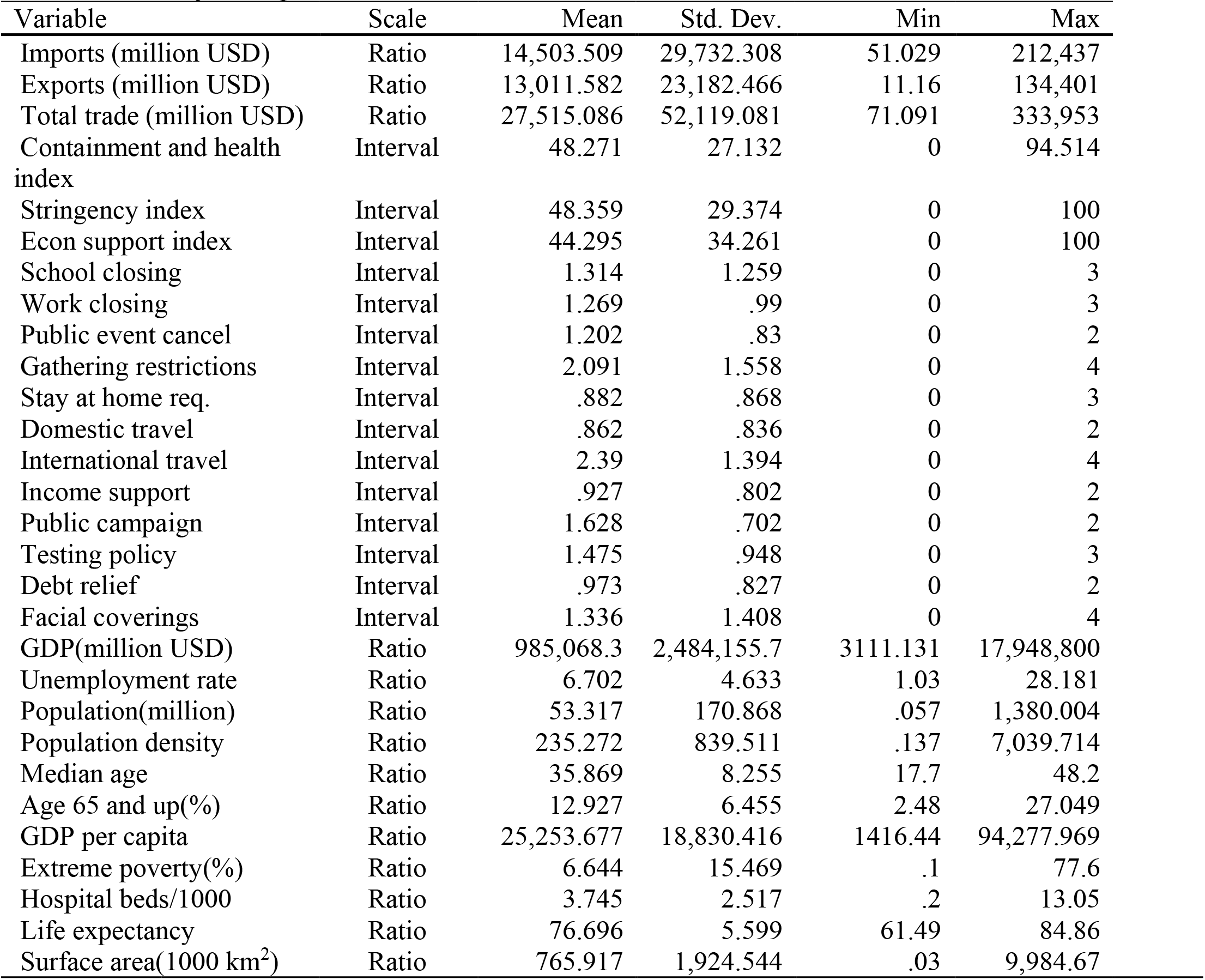
Summary descriptive statistics of the variables

It should be noted that the policy indices and specific policies are treated as interval scale in our data analysis. The policy indices and specific policies are measured by aggregating the daily ordinal scale data and convert them into monthly average data. By taking the average number of the policy indices, the data is therefore converted into interval scale. Data related to trade in terms of exports, imports and total trade values are treated as ratio scale variables. Furthermore, we include control variables to improve the robustness of our models such as GDP, population size, unemployment rate, life expectancy, extreme poverty rate and surface area.

Table 4 summarizes the random effects regression results of the types of public measures that have significant impacts on the trade flows. Three proposed models in Table 5 show the effects of policy areas on imports, exports, and total trade, respectively. Based on the results, the policy indices that have significant impact on trade values are containment and health and stringency policy areas. Containment and Health index combines lockdown restrictions and closures with healthcare-related measures such as testing policy, contact tracing, short term investment in healthcare, and investments in vaccine. SI involves the strictness of lockdown policies, which include policies that primarily target on restricting citizen’s behavior in an attempt to contain the spread of the virus. The results show that containment and health measures have significant positive effects on imports (139.809), exports (98.543) and total trade (236.643). Furthermore, stringency measures have significant negative effects on the imports (−150.912), exports (−122.452) and total trade (−272.351). This means that policies that are related to healthcare and containment are shown to be effective in containing the contraction of the trade flows during the pandemic crisis. In contrast, policies related to stringent lockdown measures tended to have negative impact on trade flows, both imports and exports, during the pandemic. Other factors shown to have significant impact on trade flows during the pandemic include countries’ GDP and land size (surface area). Countries’ with high GDP and large surface area are large countries with high trade volume. Therefore, GDP and land size are considered significant determinants of countries’ trade value. Log of GDP is shown to have significant positive impacts on imports (6,497.595), exports (5,200.253), and total trade (11,694.885). Furthermore, coefficients for surface area shows significant positive effect on imports (5.889), exports (3.554), and total trade (9.441).

**Table 4:** Impacts of COVID-19 Policy on Trade Flows: Random Effect Regression

| variable | Imports | Exports | Total Trade |
| --- | --- | --- | --- |
| Containment & health index | **139.809\*\***<br>(57.26) | **98.543\***<br>(52.954) | **236.643\*\***<br>(100.248) |
| Stringency index | **-150.912\*\*\***<br>(47.714) | **-122.452\*\*\***<br>(44.142) | **-272.351\*\*\***<br>(83.541) |
| Economic support index | 10.763<br>(12.572) | 11.092<br>(0.34) | 22.223<br>(22.008) |
| Unemployment rate | -777.593<br>(670.732) | -399.556<br>(378.868) | -1178.054<br>(1,037.953) |
| Population (million) | -11.844<br>(21.06) | -9.342<br>(11.875) | -21.182<br>(32.58) |
| Population density | 10.29<br>(24.909) | 6.813<br>(14.066) | 17.126<br>(28.544) |
| GDP (ln) | **6,497.595\*\***<br>(2745.267) | **5,200.253\*\*\***<br>(1,548.818) | **11,694.885\*\*\***<br>(4,247.419) |
| Extreme poverty (%) | 0.318<br>(250.257) | 40.516<br>(141.671) | 40.446<br>(387.638) |
| Life expectancy | 85.255<br>(764.007) | 286.035<br>(432.292) | 368.638<br>(1,182.641) |
| Surface area (1,000 km<sup>2</sup>) | **5.889\*\*\***<br>(1.714) | **3.554\*\*\***<br>(0.967) | **9.441\*\*\***<br>(2.651) |
| Constant | **-165,408.59\*\***<br>(72,463.325) | **-148,711.52\*\*\***<br>(40,972.071) | **-313,826.84\*\*\***<br>(112,155.54) |
| | $R^2 = 0.590$ | $R^2 = 0.667$ | $R^2 = 0.627$ |
|  | Num. of obs. = 301 | Num. of obs. = 301 | Num. of obs. = 301 |
| | $\chi^2 = 62.081$ | $\chi^2 = 90.887$ | $\chi^2 = 76.604$ |
| | $p\text{-value} = 0.000$ | $p\text{-value} = 0.000$ | $p\text{-value} = 0.000$ |

The standard errors are in the parentheses.
\*\*\* $p < 0.01$ , \*\* $p < 0.05$ , \* $p < 0.10$

**Table 5:** Impacts of COVID-19 specific policies on trade flows: Random Effect Regression

| Variable | Import | Export | Total Trade Value |
| --- | --- | --- | --- |
| School closing | **-1030.052\*\*\***<br>(300.668) | -280.497<br>(281.104) | **-1302.065\*\***<br>(528.059) |
| Work closing | -686.588<br>(599.411) | -849.642<br>(559.98) | -1541.119<br>(1,052.607) |
| Public event cancel | 425.844<br>(733.607) | -18.837<br>(685.301) | 394.052<br>(1,288.249) |
| Gathering restrictions | 613.517<br>(448.243) | 127.213<br>(418.091) | 730.773<br>(786.939) |
| Stay at home | **-1478.119\*\***<br>(738.149) | -715.139<br>(689.054) | **-2221.743\***<br>(1,296.074) |
| Domestic Travel | 1018.049<br>(679.659) | 149.206<br>(634.523) | 1201.222<br>(1,193.396) |
| International Travel | 328.242<br>(353.445) | 436.524<br>(329.265) | 761.31<br>(620.385) |
| Income support | 347.931<br>(562.588) | -359.797<br>(525.396) | -39.412<br>(987.885) |
| Public campaign | -426.867<br>(512.981) | -599.131<br>(479.241) | -1,017.179<br>(900.828) |
| Testing policy | -183.095<br>(456.421) | **-912.467\*\***<br>(425.836) | -1,093.608<br>(801.331) |
| Debt relief | -606.075<br>(608.186) | 779.635<br>(567.565) | 220.207<br>(1,067.825) |
| Facial covering | **726.601\*\*\***<br>(264.287) | **1,073.717\*\*\***<br>(246.742) | **1,791.447\*\*\***<br>(464.057) |
| Unemployment rate | -759.867<br>(668.57) | -377.507<br>(364.978) | -1138.299<br>(1,023.055) |
| Population (million) | -10.782<br>(21) | -9.251<br>(11.449) | -20.036<br>(32.127) |
| Population density | 10.935<br>(24.833) | 6.938<br>(13.557) | 17.912<br>(38) |
| GDP (ln) | **6,407.96\*\***<br>(2,737.11) | **5,095.9\*\*\***<br>(1,492.872) | **11,498.204\*\*\***<br>(4,187.731) |
| Extreme poverty (%) | 3.342<br>(249.435) | 47.027<br>(136.495) | 49.747<br>(381.839) |
| Life expectancy | 140.016<br>(761.775) | 385.811<br>(416.917) | 523.091<br>(1,166.176) |
| Surface area (1,000 km<sup>2</sup>) | **5.834\*\*\***<br>(1.709) | **3.64\*\*\***<br>(0.933) | **9.474\*\*\***<br>(2.616) |
| Constant | **-167061.09\*\***<br>(72,240.838) | **-153,250.43\*\*\***<br>(39,493.543) | **-319953.38\*\*\***<br>(110,570.51) |
| | $R^2 = 0.594$<br>Num. of obs. = 301<br>$\chi^2 = 97.658$<br>$p\text{-value} = 0.000$ | $R^2 = 0.670$<br>Num. of obs. = 301<br>$\chi^2 = 129.290$<br>$p\text{-value} = 0.000$ | $R^2 = 0.629$<br>Num. of obs. = 301<br>$\chi^2 = 113.181$<br>$p\text{-value} = 0.000$ |

The standard errors are in the parentheses.
\*\*\* $p < 0.01$ , \*\* $p < 0.05$ , \* $p < 0.10$

Table 5 reports the regression results of the impacts of specific policies on trade activities during COVID-19 pandemic by using Random Effects regression method. The specific policies included in the models are policies implemented by the government as intervention measures to control the spread of the COVID-19 virus. The specific policies include school closing, workplace closing, cancelling public events, restrictions on gatherings, stay at home requirements, restrictions on domestic and international travel, income support, public campaign, testing, debt relief, and facial covering policies. The results show that specific policies have different effects on imports, exports, and overall trade. In general, the specific policies that have significant effects on trade are school closing, stay-at-home, COVID-19 testing and facial covering policies. In particular, school closing and stay-at-home policies exhibit negative impacts on imports and overall trade. These policies focus on restricting people’s movement and interaction. Therefore, it is possible that these policies can hinder trade activities and reduce the import activities. Testing policy shows negative effect on exports (−912.647). Facial covering policy shows significant positive effects on imports (726.601), exports (1,073.717) and overall trade (1,791.447). Facial covering policy allows manual labor to continue working, under restricted conditions, without having to stop working completely. Therefore, it is possible that facial covering policy can have positive effect on trade.

Furthermore, GDP(log) exhibits significant positive effects on imports (6,407.96), exports (5,095.9) and total trade (11,498.204). Another control variable shown to have significant impacts on trade flows is surface area (1,000 km^2^). The coefficients for the effects of surface area on trade flows are 5.834, 3.64 and 9.474, for imports, exports and total trade, respectively. The coefficient results are graphically summarized in Fig. 7.

**Figure 7:**
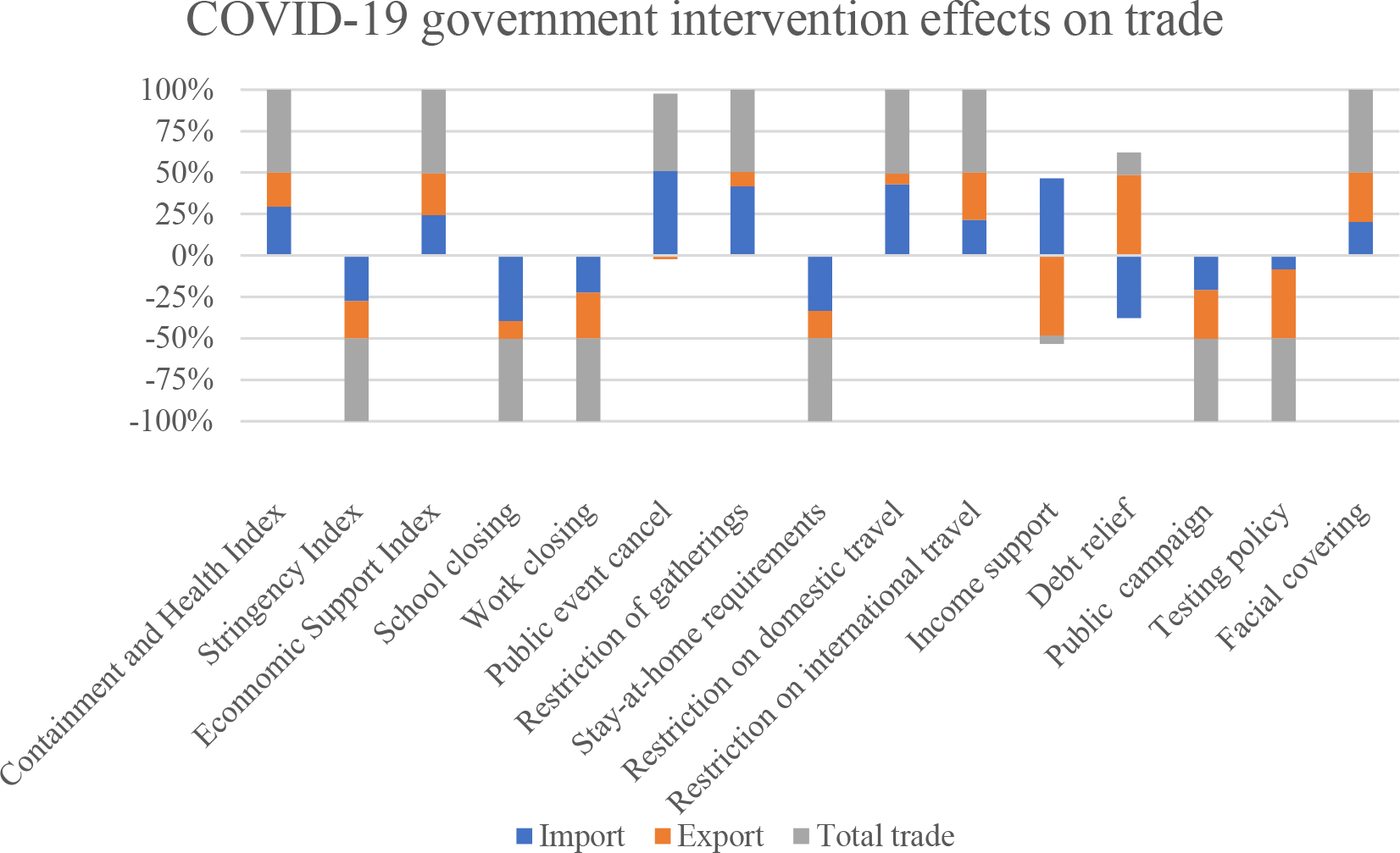
Marginal effects of COVID-19 public intervention indices and specific measures on trade flows.

## 5. Discussions

This study aimed to test three inter-related hypotheses. Our first hypothesis stated that health related policies have a positive impact on trade flows. In this context, health related policies include public awareness campaign, testing policy, contact tracing and facial covering. A relevant study conducted by Ashraf (2020b) showed that government announcements regarding public awareness programs, testing and quarantining policies, and income support packages largely result in positive stock market returns. Other studies showed that enforcing containment and healthcare policies produce benefits in terms of lowering new infections and death rates and that in turn lower death rates provide significant economic benefits in terms of saving more lives (Greenstone and Nigam 2020; Thunström et al. 2020). Our results showed that better containment and health policies represented by the containment and health index have positive impact on trade flows. Furthermore, facial covering policy was shown to have statistically significant positive impact on imports, exports and overall trade. This means that facial covering can be an effective policy in controlling the spread of COVID-19, while maintaining a continuation of economic activities in that time period.

Our second hypothesis stated that government stringent measures on closure policies, social distancing and restriction measures have negative effects on trade flows. Closure and social distancing measures in this context include school closing, workplace closing, public events cancelling, gathering restrictions, public transport closing, stay at home requirements, restrictions on internal and international travel. We assumed that restrictions on people’s movement and ability to work would greatly hinder the availability of labor force, which is very crucial to business operations. Our results are consistent with our second hypothesis, which means that closure and social distancing policies have strong negative impacts on trade flows. The results are also largely consistent with past literature. For example, Heyden and Heyden (2020), Shanaev et al. (2020) and Zaremba et al. (2020) showed that government social distancing measures are counterproductive to economic activities. A recent study concluded that containment and stringency measures have had, on average, a very large negative impact on economic activity—equivalent to a loss of about 15 percent in industrial production over a 30-day period following their implementation (Deb 2020).

Despite the direct negative impacts of stringent measures on closure and social distancing policies on trade flows, it can be argued that stringency policies can be beneficial to overall economic growth in the long run. This is because closure, movement restrictions and social distancing measures can substantially mitigate economic costs by effectively containing the spread of COVID-19 and subsequent economic effects in the long-run. Deb (2020) pointed out that stringency/containment measures are effective in mitigating some economic costs, especially the costs associated with infections and deaths from COVID-19. It was also found that among different types of containment measures, i.e. workplace closures, stay-at-home orders, and cancellations of events are effective in flattening COVID-19 related infections, but are also the costliest in terms of their impact on economic activities. Less costly containment measures, such as restrictions on international travel, can nonetheless be successful in reducing the volume of COVID-19 infections (Deb 2020).

Our third hypothesis stated that government economic support policies lead to an increase in value of trade volumes. Based on our results, we reject this hypothesis. Our findings showed that the economic support index does not have significant impact on trade flows. Specific policies related to economic support index such as debt relief and income support policies also prove to have no significant impact on trade. One possible reason is that economic support index, which we use, measures the income and debt relief support to households but not to businesses (Ashraf 2020b). However, economic support policies can have positive impact on trade if financial support were given directly to businesses related to international trade. Unfortunately, economic support index does not include financial support to businesses (Hale et al. 2020b). Thus, we can conclude that economic support policies do not have significant impact on the value of trade flows. However, economic support policies should not be entirely ignored because these policies can help alleviate the financial stress of individual citizens during the COVID-19 pandemic. Furthermore, economic support measures can help to stimulate domestic consumption and increase demand for imports in a long run. Despite tremendous government support, it would take some time for the economy to return to the pre-COVID-19 level. This is due to the fact that the income shock has a long-term impact on the private sector. In addition, businesses, especially the tourism sector, have to take time to alter their business models in order to respond to the new normal where people became more concerned regarding their health safety (BOT 2020). For this reason, economic protection for citizens through financial support to households can help the tourism sector to recover faster by increasing the number of domestic tourists to substitute for a loss of foreign tourists assuming that domestic travel is allowed. Further research on this issue could provide policy relevant evidence.

Our analysis also included the examination of the impact of specific government measures on trade. School closing policy has negative impact on imports and overall trade. School closing policy is part of the containment measures, which have direct negative impact on economic activities. Stay-at-home requirements, which is another containment measure also showed negative impact on imports and overall trade. Past study showed that containment measures have had a very large impact on economic activities, which is equivalent to a loss of 15% in industrial production over a one month period following their implementation (Deb 2020). Testing policy, which measures the implementation strictness of COVID-19 testing showed significant negative impact on exports. In contrast, Facial covering policy showed positive effects on imports, exports and overall trade value. The impact of facial covering policy on economic activities is supported by a few past studies. A recent study from Germany has shown that wearing masks is a cost-effective, less economically harmful, and democracy-compatible containment measure for COVID-19 (Mitze et al. 2020). Another study in the US suggested that facial coverings helped reduce the spread of COVID-19 without greatly disrupting economic activity if they are widely used (Knotek II et al. 2020). Therefore, based on past studies and our findings, facial covering appear to be very effective in controlling COVID-19 infection and death rates, as well as avoiding disruptions on major economic activities.

## 6. Conclusion

This study aimed to assess the impacts of COVID-19 public intervention measures on trade flows. The results show that containment and health related measures have significant positive impacts on imports, exports and overall trade values. On the other hand, stringent measures on lockdowns and social distancing measured by SI were found to have negative impact on trade. This could be due to the fact that SI focuses on the strictness of lockdown policies and tightening social distancing measures. These measures effectively reduce international trading activities by restricting labor availability and discouraging imports and exports. Economic support policies did not show a statistically significant effect. For the impact of specific policies on trade flows, school closing, stay-at-home requirements and testing policies were shown to have significant negative impact on trade flows. In contrast, facial covering policy showed significant positive impact on imports, exports and overall trade.

It should be noted that there are some limitations to this research. First of all, this paper only focuses on the impact of COVID-19 measures on a single economic activity, which is international trade. It does not take into account the potential impact of COVID-19 measures on other economic sectors that are likely to be heavily affected by the pandemic, such as manufacturing and tourism sectors. In addition, the amount of past literature related to COVID-19 is still relatively limited because the pandemic has started in 2020. Although the number of research outputs on COVID-19 is increasing rapidly, many of them are still currently under development. There is also another limitation regarding the possibility of endogeneity bias. It is possible that some policy indices and specific policies are correlated with the error terms. This means that including the specific policies into the regression models can lead to endogeneity bias. In this study, we however minimized the endogeneity bias by using Variance Inflation Factor analysis (VIF) to eliminate the variables with high VIF value. Another limitation to consider is that the data used in this study only consist of macro level data. Therefore, the results from this study did not take into account of the potential impact of COVID-19 policies at the household and individual levels.

Based on the results of this study, it is recommended that future research examine other aspects of COVID-19 government interventions, such as potential impacts on other economic sectors. In addition, as the public awareness of different sustainability issues rises, researchers can choose to explore the impact of COVID-19 on biodiversity and BioTrade. This is particularly relevant in the context of the current discussions on the forthcoming post 2020 global biodiversity framework (UNCTAD, 2021).

Furthermore, as more data become available, further research could be undertaken on a full sample of countries, or within countries and regions, which would help provide better assessment of the potential impacts of COVID-19 policies on international trade. Our results show that health-related policies have largely positive impacts on trade values. Facial covering policy in particular shows significant positive impacts on trade flows. On the other hand, stringency measures such as closure policies have negative impacts on trade flows. Therefore, governments should focus on implementing containment and health-related policies as well as maintaining less stringent measures on closure and movement restriction policies in order to minimize the loss in trade flow values amid the pandemic crisis. While we did not find a statistically significant direct effect of economic support policies on trade, such policies can be an effective tool to relieve the financial stress of households and individuals, and thus boost consumption. Further research in this area would be helpful to thoroughly understand the underlying dynamics.

## Data Availability

The data can be provided upon request.

## Acknowledgments

The authors gratefully acknowledge the valuable comments from Lisen Runsten, Camilla Blasi Foglietti, and Julia Wentworth of the World Conservation Monitoring Centre and Arlene Gonzales of the Asian Institute of Technology.

